# Retrospective multi-cohort validation of a real-world transcriptomics-guided machine learning model for treatment response prediction in breast cancer

**DOI:** 10.64898/2026.01.20.26344480

**Authors:** Hui Ren, Seth Leffel, Zheng Xu, Edwin Alphonso

## Abstract

Selection of systemic therapy for breast cancer remains largely empirical, particularly for chemotherapy, due to the lack of robust biomarkers that predict treatment response at the individual patient level. We developed Oncology CoPilot, a real-world, transcriptomics-guided machine learning (ML) decision-support model designed to integrate heterogeneous tumor gene expression data and treatment response annotations to support treatment response stratification across therapeutic classes. Oncology CoPilot was trained on a pan-cancer cohort comprising 11,414 patients across 15 cancer types and 150 systemic drug regimens derived from publicly available and published datasets. Retrospective external validation was performed using five independent breast cancer cohorts comprising 503 patients, spanning multiple molecular subtypes, transcriptomic platforms, and six commonly used treatment settings, including chemotherapy, endocrine therapy, and targeted therapy. Across the external validation cohort, the model demonstrated an overall accuracy of 72.8%, with balanced sensitivity (71.5%) and specificity (73.4%), and an ROC-AUC of 0.783. Regimen-specific analyses demonstrated stable performance for chemotherapy-based regimens (accuracy 70.1%–79.1%), highlighting the potential of transcriptomics-guided modeling to inform treatment response stratification in clinical settings where therapy selection is often empirical. For endocrine therapy, the model achieved 95.0% accuracy for tamoxifen, suggesting that transcriptomic features may capture biologically relevant estrogen-responsive and resistance-associated programs beyond receptor status alone, although this result is exploratory and based on a small sample size. In contrast, HER2-targeted therapies showed lower and more variable predictive performance, with accuracies of 66.0% for trastuzumab monotherapy and 61.9% for anthracycline–taxane chemotherapy combined with trastuzumab, likely reflecting smaller cohort sizes and the biological heterogeneity characteristic of HER2-positive disease. Overall, these findings demonstrate the feasibility of leveraging real-world transcriptomic data and machine learning to achieve generalizable treatment response stratification across diverse cohorts, platforms, and therapeutic classes, supporting the potential role of transcriptomics-guided models as complementary decision-support tools in oncology.

## Introduction

Contemporary real-world cancer treatment selection relies heavily on clinicopathologic factors and a limited set of validated biomarkers that link specific tumor alterations to approved targeted therapies or immunotherapies [1]. Despite substantial advances in precision oncology, multiple large studies and systematic reviews have consistently shown that only a subset of patients ultimately receive genomics-guided matched therapies in routine clinical practice, due to the absence of actionable alterations in many tumors, restrictive eligibility criteria, and limited access to matched treatment options [1,2]. Consequently, a substantial proportion of patients across tumor types continue to receive systemic therapies selected empirically, particularly cytotoxic chemotherapy and later-line regimens, where treatment response rates are heterogeneous and difficult to predict prospectively at the individual patient level [2,3]. These real-world limitations underscore the need for data-driven decision-support approaches capable of stratifying likely responders and non-responders beyond biomarker frameworks, especially for chemotherapy, where biologically informed guidance remains limited in current clinical practice [3].

Tumor transcriptomic profiling offers a complementary lens for therapy selection because gene expression captures dynamic biological programs and pathway activity that may not be evident from DNA alterations alone [4]. This concept has been evaluated clinically in the WINTHER trial, which used fresh-biopsy DNA sequencing or RNA-expression profiling (tumor–normal comparison) to guide therapy selection for patients with advanced cancers, illustrating how transcriptomic information can expand matching opportunities beyond DNA-only approaches [4]. In breast cancer, clinically adopted gene-expression assays demonstrate that transcriptomic measurements can meaningfully inform systemic therapy decisions and refine which patients benefit from adjuvant chemotherapy in specific settings, including prospective randomized evidence with the 21-gene recurrence score and the 70-gene signature strategy [5–7]. Beyond established assays, multiple recent studies continue to report transcriptomic signatures associated with treatment outcomes, particularly in chemotherapy response contexts such as triple-negative breast cancer, supporting the broader premise that expression patterns can encode therapy-relevant biology and may help stratify likely responders and non-responders [8–10].

Machine learning methods have advanced rapidly for drug response prediction, including models that integrate high-dimensional molecular features and leverage modern representation learning to improve predictive performance [11]. Sammut et al. demonstrated that integrating multi-omics data and tumor ecosystem features with machine-learning approaches can predict therapy response in specific clinical settings such as breast cancer, reinforcing the potential of data-driven modeling beyond single biomarkers [12]. More recently, Pal et al. reported a large-scale, transcriptomics-driven supervised learning framework that assembles multi-cohort patient gene-expression data to evaluate treatment-response prediction across multiple cancer types and therapies [13]. However, oncology machine-learning models with rigorous large-scale external validation and demonstrated clinical usability or decision-support utility remain uncommon; recent evidence syntheses indicate that many published models lack robust external validation and are rarely evaluated for real-world clinical impact [14].

To address these limitations, Oncology CoPilot was developed as a machine learning–based clinical decision-support model designed to aggregate multi-sourced transcriptomic and treatment-response data from 11,917 real-world cancer patients and to support drug selection across both cytotoxic chemotherapy regimens and targeted therapies. In this study, we report retrospective validation results of Oncology CoPilot across six common breast cancer treatment regimens, using five independent external breast cancer datasets comprising 503 patients, drawn from harmonized diverse cohorts.

## Methods

### Data collection

Publicly available pan-cancer datasets published in or before 2023 that included both clinical treatment-response data and tumor transcriptomic profiling were considered for model development. For each patient, only the first systemic drug treatment and its associated clinical response following transcriptomic profiling were included in the analysis. Subsequent lines of therapy and corresponding outcomes were excluded to reduce confounding related to treatment sequencing. Treatments involving radiation therapy were excluded; only systemic drug treatments were retained. In total, 199 pan-cancer datasets from published sources were included in the training cohort, comprising 153 datasets from GEO, 29 datasets from TCGA, 11 datasets from ArrayExpress, 5 datasets from the Genomic Data Commons (GDC), and 1 additional dataset reported in peer-reviewed publications. Collectively, these datasets included 11,917 patients across 15 major cancer types and 150 commonly used systemic drug regimens. Breast cancer represented the largest cancer type comprising 4,844 cases.

### Clinical and drug data curation

Clinical and treatment data were curated from heterogeneous sources and provided in multiple formats, including CSV, TSV, JSON, XML, and XLSX files, as well as supplementary tables embedded within published manuscripts. Clinical variables collected included de-identified patient identifiers, cancer type, treatment regimens, treatment line, treatment start and end dates (when available), and reported treatment response outcomes. Drug information underwent manual curation and normalization to harmonize inconsistent naming conventions across datasets. Drug names were standardized to generic names, and combination regimens were decomposed into constituent agents using curated drug dictionaries and reference mappings. Treatment course information, including treatment sequencing, duration, and temporal alignment relative to transcriptomic profiling, was extracted when available and used to define eligible treatment–response pairs. Only systemic drug treatments were retained for analysis. To ensure consistency across datasets, clinical response annotations derived from diverse criteria (e.g., RECIST-based assessments, pathological response, or study-specific response definitions) were harmonized into unified 2 outcome categories (response vs. non-response). Complete response (CR) and partial response (PR) were classified as responders, while stable disease (SD), progressive disease (PD), and non-response were classified as non-responders, consistent with common retrospective oncology analyses. Records with ambiguous, missing, or conflicting treatment or outcome annotations were excluded. All curation steps were performed on de-identified data in accordance with the governance policies of the original data providers.

### Transcriptomic data processing

Tumor whole transcriptomic RNA sequencing data were obtained from multiple public repositories and published studies and included both raw sequencing data and processed gene expression matrices, depending on dataset availability. For datasets with available raw FASTQ files, RNA sequencing data were processed using a standardized alignment and quantification workflow. Briefly, raw reads were aligned to the human reference genome (GRCh38) using the STAR aligner with a two-pass alignment strategy, consistent with previously published pipelines used by the Genomic Data Commons (GDC) [15]. Gene-level read counts were generated, and transcript abundance was subsequently normalized to transcripts per million (TPM) to facilitate cross-sample and cross-dataset comparability. For datasets in which raw FASTQ files were not available, published gene-level raw count matrices or normalized expression values were used as provided. When raw gene counts were available, TPM values were computed using gene length annotations consistent with the reference genome. For datasets distributed only as normalized TPM expression values, the original processed values were retained. For microarray-based transcriptomic datasets, normalized expression matrices provided by the original studies were used. To mitigate systematic technical variability across studies and platforms, batch effects were adjusted using the ComBat method, with batch variables defined by study or dataset of origin. Breast cancer molecular subtypes were assigned using the previously published PAM50 intrinsic subtype classifier, which categorizes tumors into Basal, Luminal A, Luminal B, HER2-enriched, and Normal-like based on the expression patterns of 50 classifier genes [16].

### Machine learning models

The Oncology CoPilot modeling framework integrates multi-level biological and pharmacological feature representations derived from tumor transcriptomic profiles and drug-related information. Feature construction was guided by established biological knowledge and prior empirical performance in drug response modeling studies. Tumor-intrinsic features include expression patterns of cancer driver genes, transcriptional signatures capturing coordinated biological processes, and tumor subtype–associated expression patterns reflecting known molecular heterogeneity. Drug-related features incorporate drug target annotations and mechanism-of-action (MoA) representations to capture shared molecular properties and biological functions across chemotherapies, targeted agents, endocrine therapies, immunotherapies, and combination regimens. Together, these curated features enable the model to learn differential treatment sensitivity across heterogeneous tumor profiles and therapy classes. Multiple supervised machine learning algorithms, including random forest, gradient boosting machines, deep learning models, and other approaches previously shown to perform well in drug response prediction tasks [17–21], were trained and integrated using an ensemble strategy. Five-fold cross-validation (5-fold CV) was used during model development to optimize performance and assess internal generalization. This ensemble design is intended to enhance robustness and generalizability across diverse cancer types, therapeutic classes, and heterogeneous data quality profiles. Model training was performed using retrospective treatment–response pairs. Detailed algorithmic architecture, feature engineering, and training and optimization procedures are intentionally abstracted in this manuscript due to ongoing intellectual property protection. Accordingly, this study focuses on retrospective validation results and predictive performance, rather than methodological novelty.

### Training & validation evaluation

The model was trained on a pan-cancer cohort comprising 11,414 patients spanning 15 major cancer types and 150 commonly used systemic treatment regimens. Among these, breast cancer represented the largest cancer type, with 4,341 cases, and was therefore selected for focused external validation to enable meaningful evaluation. Five independent breast cancer datasets, comprising a total of 503 patients, were used for external validation. These datasets included GSE163882, GSE32646, GSE22093, GSE44272, and GSE82171 and were used exclusively for retrospective validation, with no overlap with the training data. The validation cohorts included both RNA sequencing–based and microarray-based gene expression profiles, enabling assessment of model robustness across distinct transcriptomic platforms. These five datasets were selected because their treatment regimens were fully represented within the training cohort, ensuring that validation focused on generalization across independent patient cohorts and data sources, rather than extrapolation to previously unseen drug regimens. This design enabled evaluation of predictive performance under clinically realistic conditions without introducing confounding from out-of-distribution therapies, while preserving the full size and diversity of the training cohort. It further allowed assessment of model generalizability across diverse treatment classes, including chemotherapy combinations, targeted therapy, and endocrine therapy, as well as across independent cohorts, heterogeneous transcriptomic platforms, and real-world clinical regimens.

### Computational Environment and Software

Data processing and analysis were performed using a combination of bash shell scripting, R, and Python. Bash scripts were used to orchestrate large-scale file handling and reproducible execution of transcriptomic preprocessing workflows. Clinical data harmonization and gene expression matrix processing including normalization, batch adjustment and subtype PAM50 classification were performed primarily in R. Machine learning model development, training, and evaluation were implemented in Python.

### Ethics and Data Governance

All data used in this study were obtained from publicly available or published sources. All datasets were de-identified prior to analysis in accordance with the original data providers’ governance policies, and no direct patient contact or clinical intervention was performed. As this study involved secondary analysis of de-identified data, formal institutional review board (IRB) approval was not required. This work is intended to provide insights into tumor biology derived from transcriptomic profiles and to support clinical reasoning around treatment selection; it does not provide medical advice, replace clinician judgment, or make treatment recommendations. Model performance may be influenced by cohort composition and biases inherent in retrospective real-world datasets. The datasets analyzed are publicly available or derived from published sources, with access governed by the original data providers’ terms and applicable data use agreements.

## Results

### Study Workflow Overview

An overview of the study workflow is shown in Figure 1. Briefly, transcriptomic and clinical treatment response data were downloaded and harmonized from multiple public and published sources, yielding a pan-cancer study cohort of 11,917 patients. The final dataset comprised 11,917 unique patients, including 11,414 patients in the pan-cancer training cohort and 503 patients reserved exclusively for external breast cancer validation. Clinical data curation was performed to standardize treatment regimens, treatment timing, and response annotations, ensuring consistency across heterogeneous datasets. Transcriptomic data engineering was conducted on both RNA-sequencing and microarray datasets, enabling integration across platforms. In parallel, drug-related knowledge representations were constructed to capture relevant therapeutic characteristics for systemic treatments included in the study. These components were jointly incorporated into a machine learning modeling framework designed to learn associations between tumor transcriptomic profiles, treatment features, and clinical response outcomes. Model training was performed on the pan-cancer cohort spanning 15 major cancer types and 150 commonly used systemic drug regimens. Given that breast cancer represented the largest disease group in the training data, external validation was focused on this indication. Model performance was evaluated using five independent breast cancer datasets, comprising a total of 503 patients. These validation cohorts included both RNA-sequencing–based and microarray-based transcriptomic profiles, enabling assessment of model robustness across diverse data sources and experimental platforms. Area Under the Receiver Operating Characteristic Curve (ROC-AUC) and confusion matrix were used for clinical evaluation. True positives (TP), true negatives (TN), false positives (FP), and false negatives (FN) were calculated for the external validation cohort. Based on these values, standard classification performance metrics, including sensitivity, specificity, and overall accuracy, were computed to evaluate model performance.

**Figure 1.**
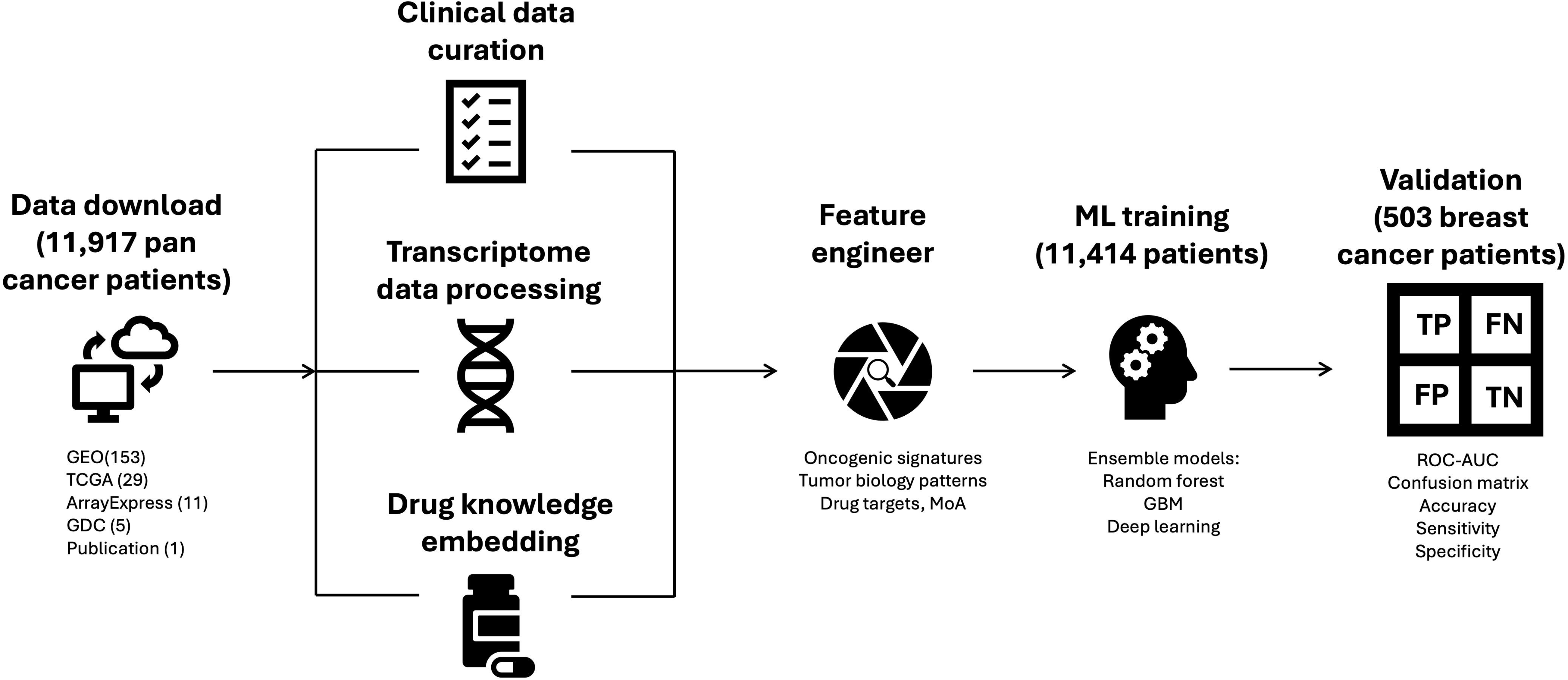
Study workflow overview. Transcriptomic and clinical treatment response data from 11,917 real-world cancer patients were curated, harmonized, and integrated from multiple publicly available and published data sources. Tumor transcriptomic data underwent standardized processing and normalization, and drug-related knowledge was incorporated through high-level feature representation. These inputs were integrated within a machine learning modeling framework trained on a pan-cancer cohort spanning 15 cancer types and 150 systemic treatment regimens. External validation was conducted using five independent breast cancer datasets comprising 503 patients, representing diverse molecular subtypes, transcriptomic platforms, and treatment classes. Model evaluation included assessment of discriminative performance using ROC-AUC, as well as classification performance based on confusion matrix–derived metrics, including accuracy, sensitivity, and specificity.

### Cohort Overview

The aggregated study cohort comprised 11,917 patients drawn from 199 published datasets (Figure 2A) from multiple data sources. The cohort was clinically and biologically diverse, encompassing a broad range of age distributions, sex, disease stages, and racial and ethnic backgrounds, as reported in the original data sources. Tumor transcriptomic data were generated using multiple technology platforms, including 4,301 RNA-sequencing samples and 7,616 microarray-based assays (Figure 2B), reflecting heterogeneous sample preparation and processing protocols consistent with real-world oncology datasets. Across the full cohort, treatment response outcomes were relatively balanced, with 6,080 non-responders and 5,837 responders, supporting supervised modeling without extreme class imbalance. Patients represented 15 major cancer types (Figure 2C), with substantial representation across several indications. Seven cancer types contributed more than 500 cases each, including breast cancer, leukemia, colorectal cancer, lung cancer, bone marrow cancer, melanoma, and ovarian cancer. Breast cancer constituted the largest disease group, accounting for approximately 38% of the total cohort. A total of 150 commonly used systemic drug regimens were represented across the dataset, spanning cytotoxic chemotherapy, targeted therapy, immunotherapy, endocrine therapy, and combination regimens (Figure 2D). Of these, 103 regimens (68.7%) were combination therapies, 21 were single-agent targeted therapies, 16 were single-agent chemotherapies, 6 were immunotherapies, and 4 were endocrine therapies. Collectively, these regimens reflect treatment strategies routinely used in real-world oncology practice and enable broad evaluation of treatment response prediction across diverse therapeutic classes.

**Figure 2.**
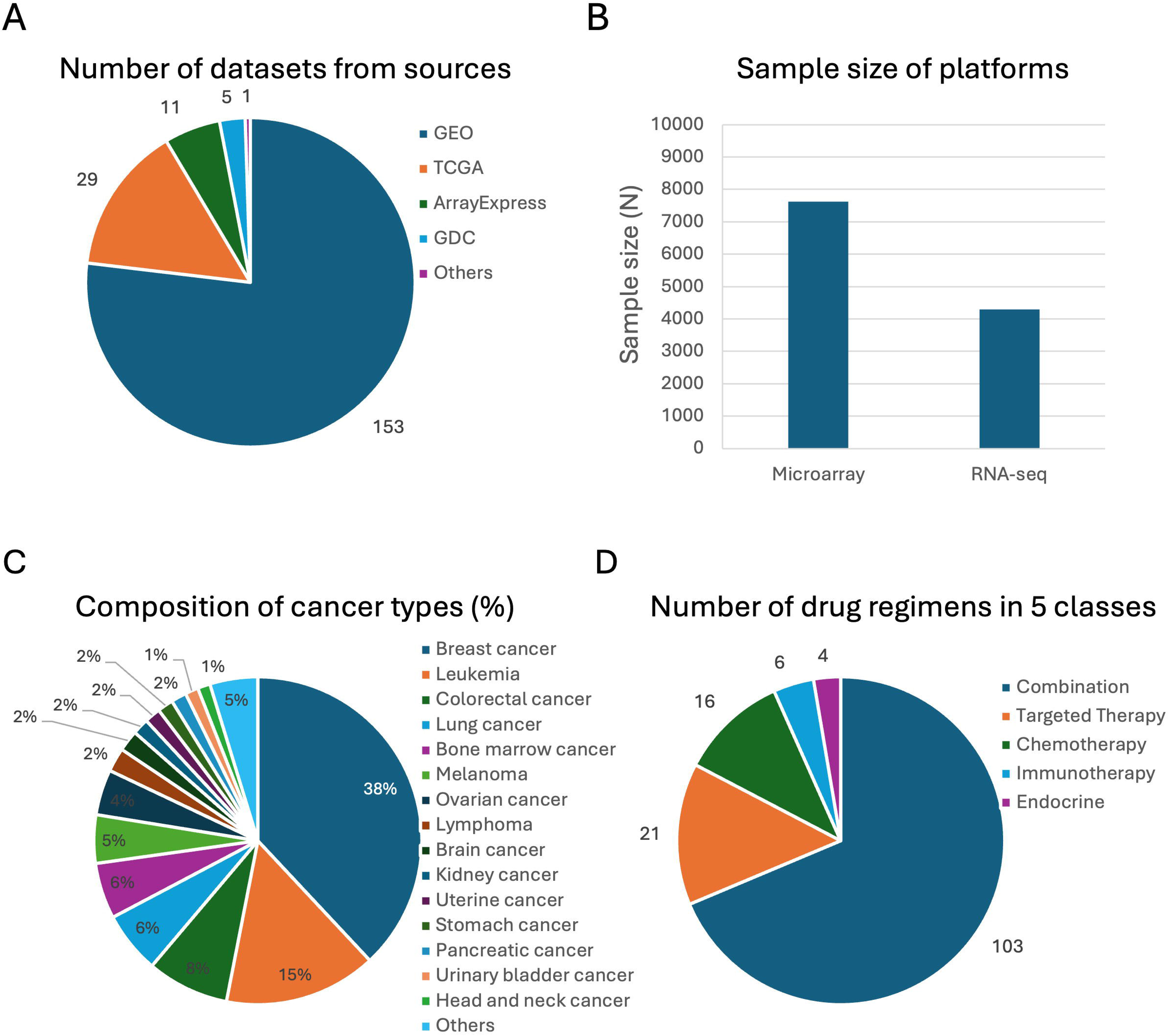
Aggregated data cohort overview. (A) Distribution of datasets across five primary data sources, including GEO, TCGA, ArrayExpress, GDC, and other published sources, illustrating the multi-source nature of the aggregated cohort. (B) Distribution of patient cases by transcriptomic data type, showing contributions from microarray-based and RNA-sequencing–based gene expression profiles. (C) Cancer type composition of the aggregated cohort, represented as percentages across 15 major cancer types, with breast cancer constituting the largest disease group. (D) Distribution of 150 systemic drug regimens across five therapeutic classes, including combination therapies, targeted therapies, chemotherapy, immunotherapy, and endocrine therapy, reflecting the diversity of real-world treatment strategies represented in the study.

### Oncology CoPilot demonstrates generalizable predictive performance across treatment settings

Five independent external breast cancer datasets comprising 503 patients, representing approximately 10.4% of all breast cancer cases in the aggregated cohort, were used for retrospective validation. These patients encompassed five molecular subtypes defined by the PAM50 classifier (Figure 3A), reflecting biological heterogeneity within the validation population. Basal accounted for 139 cases, HER2-enriched tumors for 102 cases, Luminal A tumors for 87 cases, Luminal B tumors for 154 cases and Normal-like for 21 cases. Transcriptomic profiling in the validation cohorts included both RNA sequencing (n = 218 from 1 dataset) and microarray-based expression data (n = 285 from 4 datasets) (Figure 3B), enabling assessment of model performance across distinct transcriptomic platforms. The validation cohorts spanned six commonly used breast cancer treatment settings, which represent diverse therapeutic classes, including chemotherapy, endocrine therapy, targeted therapy, and combination regimens, and reflect treatment strategies routinely used in real-world breast cancer care.

**Figure 3.**
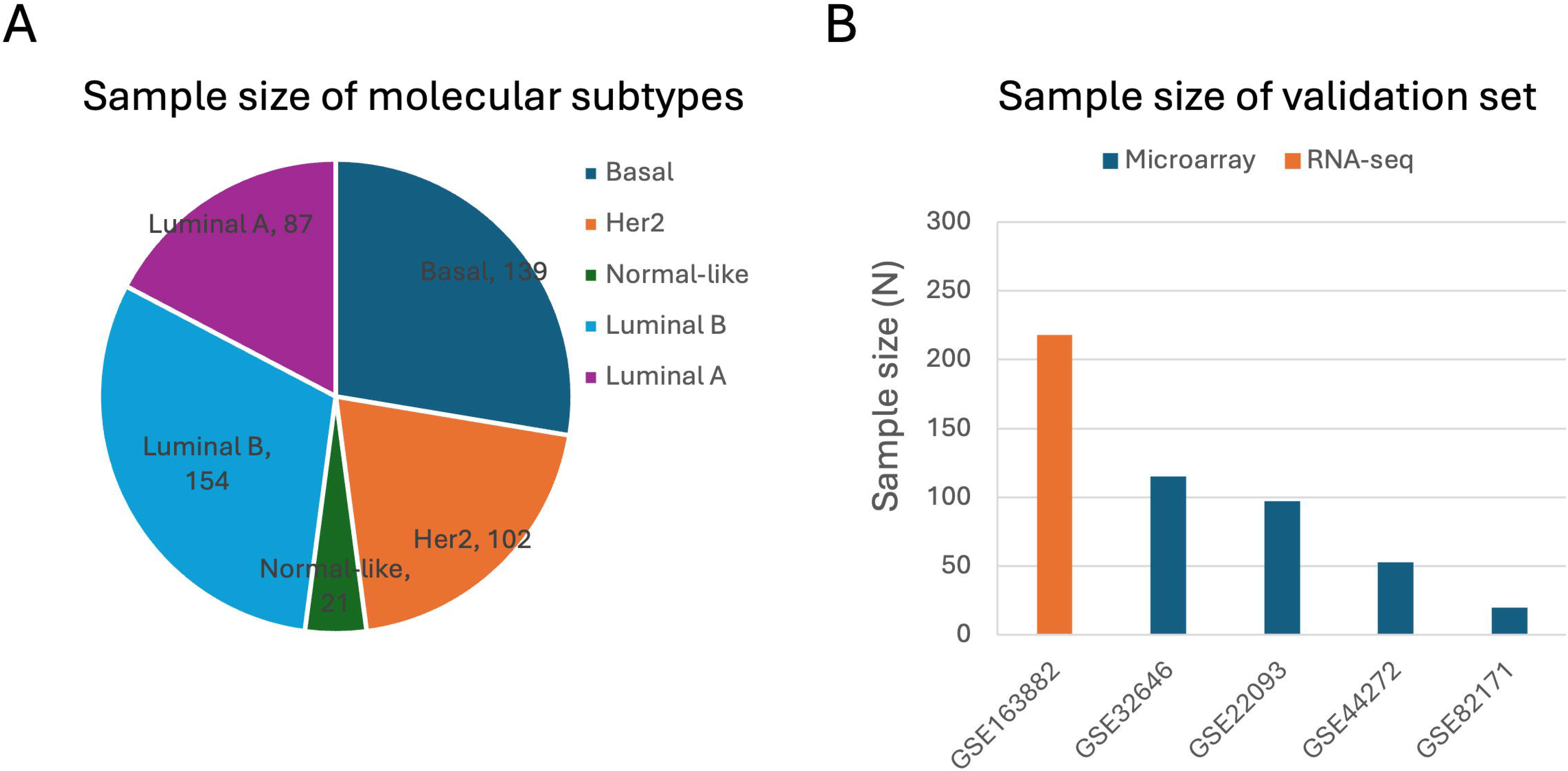
Characteristics of the external breast cancer validation cohorts. (A) Distribution of breast cancer molecular subtypes in the external validation cohort, classified using the PAM50 scheme, including Basal, HER2-enriched, Luminal A, Luminal B and Normal-like subtypes. (B) Distribution of validation cases by transcriptomic data platform across the five independent breast cancer datasets, showing contributions from RNA-sequencing and microarray-based gene expression profiles.

Across heterogeneous external cohorts and treatment settings, ROC-AUC analysis demonstrated robust and consistent discriminative performance of the stacked ensemble model.

During five-fold cross-validation, the model achieved an ROC-AUC of 0.815, indicating strong internal discrimination. When evaluated on a fully independent external validation cohort that was not used during model training or tuning, performance remained high, with an ROC-AUC of 0.783. The similarity in ROC curve profiles between cross-validation and external validation, together with the modest reduction in ROC-AUC, suggests limited overfitting and good generalizability across heterogeneous real-world samples (Figure 4A). Collectively, these results indicate that Oncology CoPilot maintains stable and reproducible predictive performance when transitioning from internal validation to independent, real-world datasets.

**Figure 4.**
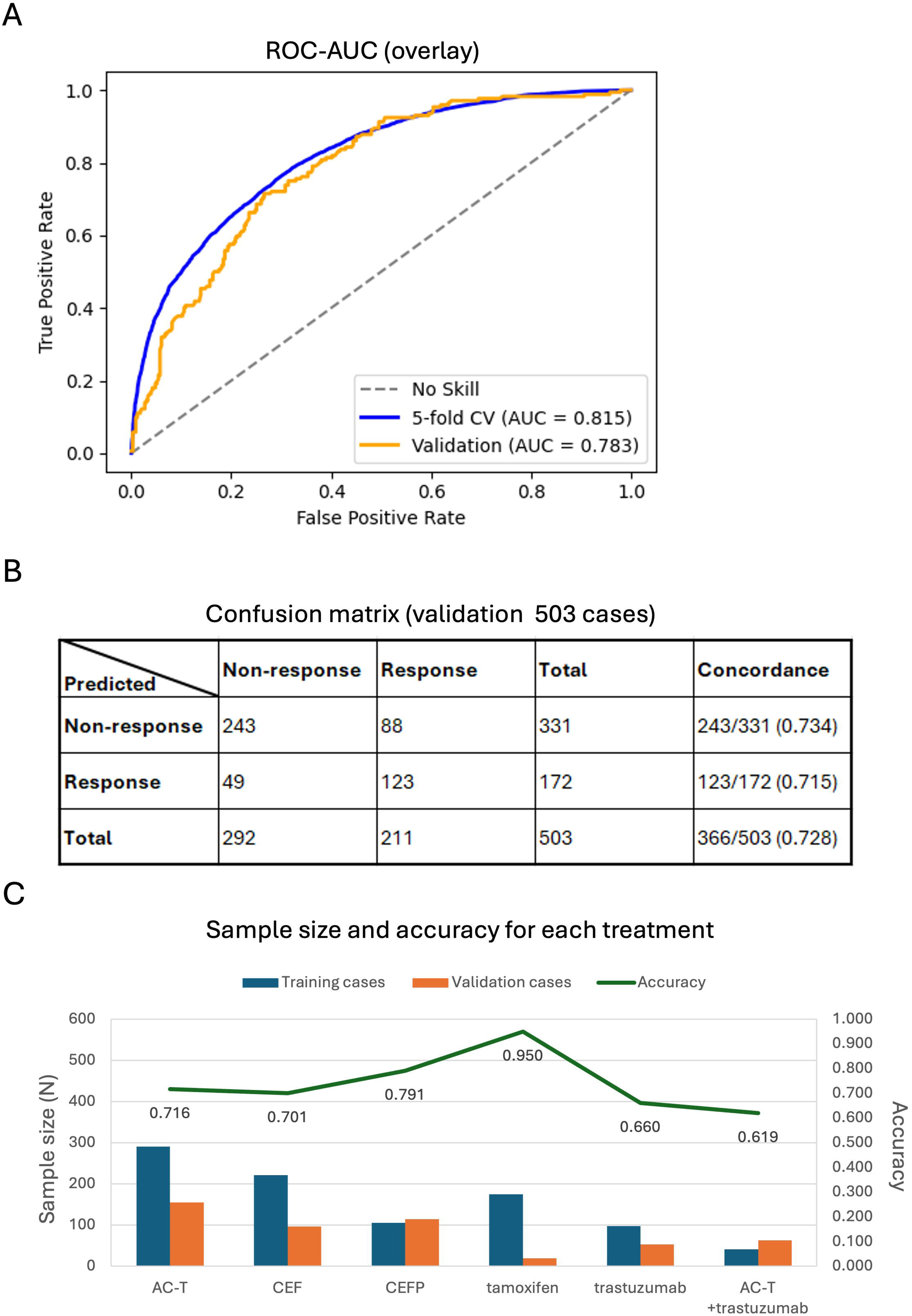
Model performance on the external breast cancer validation cohort. (A) ROC curves comparing model performance under five-fold cross-validation (blue) and external validation (orange). The ML model achieved an ROC-AUC of 0.815 during five-fold cross-validation on the training cohort and an ROC-AUC of 0.783 when evaluated on an independent external validation cohort. The diagonal dashed line represents no-discrimination (random chance). (B) Confusion matrix summarizing classification performance for 503 external validation cases, showing the distribution of predicted versus observed treatment response categories. The table reports the number of true positives, true negatives, false positives, and false negatives, along with class-specific correct match rates and overall accuracy. (C) Regimen-specific prediction performance, illustrating the number of cases used for model training and external validation for each breast cancer treatment setting, alongside corresponding prediction accuracy. Treatment regimens include AC-T, CEF, CEFP, tamoxifen, trastuzumab, and AC-T plus trastuzumab, highlighting variation in predictive performance across therapeutic classes and cohort sizes.

Classification performance on the external breast cancer validation cohort was evaluated using a confusion matrix (Figure 4B). Among 331 non-responder cases, the model correctly classified 243 patients to the non-responder category, corresponding to a specificity of 73.4%. Among 172 responder cases, 123 patients were correctly classified as responders, yielding a sensitivity of 71.5%. Overall, 366 of 503 patients were correctly classified, corresponding to an overall accuracy of 72.8% across the external validation cohort. The comparable sensitivity and specificity indicate balanced classification performance between responder and non-responder groups, without a strong bias toward either outcome class.

These validation results suggest that Oncology CoPilot maintains stable and generalizable predictive performance across heterogeneous breast cancer molecular subtypes, transcriptomic data platforms, and therapeutic classes in real-world settings.

### Oncology CoPilot shows regimen-specific variation in predictive performance

Prediction performance was further evaluated at the individual drug regimen level across the six breast cancer treatment settings included in external validation. These settings comprised AC-T (cyclophosphamide + doxorubicin + paclitaxel), CEF (cyclophosphamide + epirubicin + fluorouracil), CEFP (cyclophosphamide + epirubicin + fluorouracil + paclitaxel), tamoxifen, trastuzumab, and AC-T plus trastuzumab (cyclophosphamide + doxorubicin + paclitaxel + trastuzumab). Training and validation sample sizes for each regimen, along with corresponding prediction accuracies, are summarized in Figure 4C. Among combination chemotherapy regimens, the model achieved an accuracy of 71.6% for AC-T (111 correct predictions out of 155 validation cases) and 70.1% for CEF (68 of 97). For the four-drug combination CEFP, prediction accuracy reached 79.1% (91 of 115), representing the highest accuracy observed among multi-agent chemotherapy regimens. Notably, these chemotherapy-based regimens were supported by larger training and validation sample sizes (approximately >100 cases) and demonstrated more stable and consistently high predictive performance, consistent with improved model learning in well-represented treatment settings. For endocrine therapy, the model achieved an accuracy of 95.0% for tamoxifen (19 correct predictions out of 20 validation cases), although this result requires confirmation in larger cohorts. In contrast, predictive performance for trastuzumab-based therapies was comparatively lower. The model achieved 66.0% accuracy for trastuzumab monotherapy (35 of 53 validation cases) and 61.9% accuracy for the combination regimen AC-T plus trastuzumab (39 of 63). Notably, both trastuzumab-containing regimens were represented by smaller training and validation cohorts (<100 cases) relative to chemotherapy-only regimens, which may contribute to increased performance variability and is consistent with the biological and clinical heterogeneity characteristic of HER2-positive breast cancer.

Overall, regimen-specific analyses demonstrated meaningful predictive performance across diverse systemic treatment classes, including chemotherapy, endocrine therapy, targeted therapy, and combination regimens. At the same time, observed differences in accuracy highlight the influence of treatment complexity, biological heterogeneity, and cohort size on predictive performance in real-world datasets.

## Discussion

Anthracycline- and taxane-based chemotherapy regimens, including AC-T (cyclophosphamide + doxorubicin + paclitaxel), CEF (cyclophosphamide + epirubicin + fluorouracil), and CEFP (cyclophosphamide + epirubicin + fluorouracil + paclitaxel), remain standard systemic treatments for breast cancer across neoadjuvant and adjuvant settings. In routine clinical practice, selection among these regimens is primarily guided by clinicopathologic factors rather than by validated predictive biomarkers of chemotherapy sensitivity. Across neoadjuvant studies, treatment efficacy is most commonly assessed using pathologic complete response (pCR), which remains modest overall and strongly subtype-dependent. Large randomized trials and pooled analyses report pCR rates of approximately 10–15% for anthracycline-only regimens such as CEF, while the addition of a taxane increases pCR rates to approximately 20–30% overall, with higher rates observed in triple-negative disease and lower rates in hormone receptor–positive subtypes [22–24]. For example, in NSABP B-27, the addition of a taxane after AC increased pCR from 13.7% to 26.1%, illustrating both the benefit and the persistent heterogeneity of chemotherapy response [23]. In contrast to these population-level response proportions, Oncology CoPilot demonstrated patient-level predictive accuracy of 71.6% for AC-T, 70.1% for CEF, and 79.1% for CEFP in external real-world validation. Although these accuracy values do not represent treatment efficacy and are not directly comparable to pCR rates, they reflect the model’s ability to discriminate responders from non-responders within heterogeneous cohorts. Notably, CEFP as the most intensive multi-agent regimen evaluated achieved the highest predictive accuracy, aligning with the notion that response to combination chemotherapy is driven by global tumor biological programs such as proliferation, DNA damage response, and cell-cycle activity, which are well captured by transcriptomic profiles. The relatively stable performance observed across chemotherapy regimens is further supported by larger training and validation sample sizes (approximately >100 cases) compared with targeted therapy settings. Taken together, these comparisons highlight a critical distinction between clinical response prevalence (e.g., pCR rates of ∼10–30%) and predictive discrimination (Oncology CoPilot accuracy of ∼70–80%). While traditional clinical studies quantify how many patients respond to chemotherapy, Oncology CoPilot addresses a complementary and unmet need: identifying which patients are more likely to respond within real-world populations where chemotherapy selection remains largely empirical. These findings suggest that transcriptomics-guided machine learning may provide biologically informed stratification to refine chemotherapy decision support, particularly in treatment contexts where validated predictive biomarkers are lacking.

Tamoxifen is a foundational endocrine therapy for estrogen receptor–positive (ER+) breast cancer, used in both adjuvant and advanced disease settings. Tamoxifen demonstrates modest but clinically meaningful activity that varies by disease stage and prior endocrine exposure. In the neoadjuvant setting for ER-positive breast cancer, clinical response rates of approximately 30–50% have been reported, although pathologic complete response remains rare [25,26]. In metastatic disease, tamoxifen achieves objective response rates of approximately 30–40% when used as first-line endocrine therapy in endocrine-naïve patients [27, 28], while later-line use following prior endocrine exposure is associated with lower response rates of approximately 10–20%, reflecting the emergence of endocrine resistance mechanisms [29]. Despite ER positivity being required for treatment eligibility, a substantial proportion of ER+ tumors exhibit de novo or acquired endocrine resistance, underscoring the limitations of single-biomarker selection. In contrast to these population-level response rates, Oncology CoPilot achieved a regimen-level prediction accuracy of 95.0% for tamoxifen in external validation. This accuracy does not represent treatment efficacy and should be interpreted cautiously given the limited validation sample size (n = 20). Rather, it reflects the model’s ability to discriminate responders from non-responders within a labeled cohort. The high observed accuracy is biologically plausible, as transcriptomic profiles capture estrogen-responsive transcriptional programs and resistance-associated gene expression states beyond ER status alone. Larger endocrine-treated validation cohorts will be required to confirm the stability and generalizability of this finding.

Trastuzumab targets the HER2 receptor, which is amplified or overexpressed in approximately 20-30% of breast cancers. HER2 positivity is a necessary biomarker for trastuzumab eligibility; however, HER2 amplification alone is an incomplete predictor of therapeutic response. In metastatic breast cancer, clinical trials and pooled analyses report objective response rates to trastuzumab monotherapy of approximately 20–35%, even among biomarker-selected HER2-positive patients, reflecting substantial biological heterogeneity and resistance mechanisms [30–32]. Within this context, Oncology CoPilot achieved a prediction accuracy of 66.0% for trastuzumab monotherapy in external validation. While lower than the accuracy observed for chemotherapy regimens, this performance should be interpreted in light of smaller training and validation sample sizes (<100 cases) and the known complexity of HER2-driven disease. Response to HER2-targeted therapy is influenced by factors such as downstream signaling alterations (e.g., PI3K/AKT), immune-mediated effects, tumor heterogeneity, and prior treatment exposure, many of which may not be fully captured by baseline bulk transcriptomic profiles alone. Thus, the observed accuracy reflects the challenges of predicting response to targeted therapy using transcriptomics in isolation, while still suggesting complementary discriminatory signal beyond HER2 status.

The combination of anthracycline–taxane chemotherapy with trastuzumab represents a standard-of-care regimen for HER2-positive breast cancer, particularly in the neoadjuvant setting. Clinical trials consistently demonstrate that adding trastuzumab to chemotherapy substantially improves outcomes compared with chemotherapy alone. Early neoadjuvant studies with small sample size showed that trastuzumab combined with anthracycline- and taxane-based chemotherapy could achieve pathologic complete response (pCR) rates exceeding 60% in selected HER2-positive populations, compared with approximately 25–30% with chemotherapy alone, establishing the biological impact of HER2-targeted therapy in this setting [33]. Subsequent larger trials and real-world experience have reported more typical pCR rates in the range of approximately 30–40% for chemotherapy plus trastuzumab, with substantial variation according to hormone receptor status, regimen composition, and patient selection [34,35]. Despite these high population-level response rates, a meaningful subset of HER2-positive patients still fail to achieve pCR, highlighting persistent inter-patient heterogeneity. In external validation, Oncology CoPilot achieved a prediction accuracy of 61.9% for the AC-T plus trastuzumab regimen. As with trastuzumab monotherapy, this accuracy metric reflects patient-level discrimination rather than absolute treatment efficacy and should be interpreted cautiously given the limited cohort sizes for trastuzumab-containing regimens. The comparatively lower accuracy relative to chemotherapy-only regimens may reflect the increasing influence of non-transcriptional determinants of response, including immune engagement, receptor signaling dynamics, and treatment sequencing effects. These findings suggest that while transcriptomics-guided modeling provides informative signal, multimodal integration may be particularly important for optimizing prediction of response to targeted combination therapies.

Several limitations of this study should be acknowledged. First, sample sizes for certain regimens, particularly endocrine and HER2-targeted therapies, were limited, which may contribute to variability in performance estimates. Second, the aggregated datasets were generated using diverse transcriptomic platforms, sample preparation methods, and processing pipelines, introducing technical heterogeneity despite batch correction and harmonization efforts. Third, real-world clinical annotations, including treatment response definitions and timing, were derived from non-standardized sources and may contain noise or incompleteness inherent to retrospective datasets.

Future work will focus on expanding both training and validation datasets to encompass common cancer types and widely used systemic treatment regimens through multi-institutional collaboration and data-sharing efforts. Increasing representation across standard-of-care chemotherapies, targeted therapies, endocrine treatments, and immunotherapy regimens will enable more comprehensive evaluation of model performance and improve generalizability across real-world clinical scenarios. In parallel, incorporation of larger and more diverse external validation cohorts will strengthen assessment of robustness across cancer subtypes, molecular contexts, and transcriptomic platforms. Importantly, prospective, case-based, and real-world clinical evaluations will be necessary to rigorously assess clinical usability, interpretability, and decision-support value of transcriptomics-guided models within routine oncology workflows.

## Conclusion

Taken together, these findings demonstrate that a real-world transcriptomics-guided machine learning model can achieve generalizable and reproducible predictive performance across multiple treatment regimens, therapeutic classes, transcriptomic platforms, and heterogeneous patient cohorts. By leveraging real-world tumor transcriptomic data, Oncology CoPilot enables treatment response stratification grounded in tumor biology, addressing a critical gap in current oncology practice where therapy selection—particularly for chemotherapy—remains largely empirical.

## Data Availability

All data used in this study were obtained from publicly available repositories and published sources. The original datasets are publicly accessible from the Gene Expression Omnibus (GEO), The Cancer Genome Atlas (TCGA), ArrayExpress, and the Genomic Data Commons (GDC), as well as from the supplementary materials of the corresponding published studies.

## Acknowledgements

The authors acknowledge the use of publicly available data resources, including GEO, TCGA, ArrayExpress, GDC, CTRDB as well as curated knowledge bases such as DrugBank, KEGG, PharmGKB and TTD, which were essential for data integration and analysis in this study.

We are grateful to Dr. Eric Lau (Knight Cancer Institute at Oregon Health and Science University) for valuable scientific insights and discussions, and to Dr. Andrew Ko (University of California, San Francisco) and Dr. Jennifer Valerin (University of California, Irvine) for their clinical perspectives and guidance, which greatly informed the interpretation of results and the framing of this work.

## Author contributions

**H.R.** led the conception and design of the study, defined the scientific objectives, oversaw data integration and modeling strategy, interpreted results, and drafted the manuscript. **S.L.** contributed to workflow development, data processing pipelines, software engineering, and system integration supporting model development and evaluation. **Z.X.** contributed to product management–related activities, including requirements definition, coordination of data and software workflows, usability considerations, and alignment of modeling outputs with clinical and translational objectives. **E.A.** provided executive leadership and strategic oversight, contributed to study direction and translational positioning, and supported interpretation of results in the context of real-world clinical and commercialization considerations.

## References

1. Mateo, J., Chakravarty, D., Dienstmann, R., Jezdic, S., Gonzalez-Perez, A., Lopez-Bigas, N., Ng, C. K. Y., Bedard, P. L., Tortora, G., Douillard, J. Y., van Allen, E. M., Schultz, N., Swanton, C., André, F., & Pusztai, L. (2018). A framework to rank genomic alterations as targets for cancer precision medicine: The ESMO Scale for Clinical Actionability of molecular Targets (ESCAT). Annals of Oncology, 29(9). 10.1093/annonc/mdy263

2. Tannock, I. F., & Hickman, J. A. (2016). Limits to Personalized Cancer Medicine. New England Journal of Medicine, 375(13). 10.1056/nejmsb1607705

3. Schilsky, R. L. (2010). Personalized medicine in oncology: The future is now. In Nature Reviews Drug Discovery (Vol. 9, Issue 5). 10.1038/nrd3181

4. Rodon, J., Soria, J. C., Berger, R., Miller, W. H., Rubin, E., Kugel, A., Tsimberidou, A., Saintigny, P., Ackerstein, A., Braña, I., Loriot, Y., Afshar, M., Miller, V., Wunder, F., Bresson, C., Martini, J. F., Raynaud, J., Mendelsohn, J., Batist, G., … Kurzrock, R. (2019). Genomic and transcriptomic profiling expands precision cancer medicine: the WINTHER trial. Nature Medicine, 25(5). 10.1038/s41591-019-0424-4

5. Sparano, J. A., Gray, R. J., Makower, D. F., Pritchard, K. I., Albain, K. S., Hayes, D. F., Geyer, C. E., Dees, E. C., Goetz, M. P., Olson, J. A., Lively, T., Badve, S. S., Saphner, T. J., Wagner, L. I., Whelan, T. J., Ellis, M. J., Paik, S., Wood, W. C., Ravdin, P. M., … Sledge, G. W. (2018). Adjuvant Chemotherapy Guided by a 21-Gene Expression Assay in Breast Cancer. New England Journal of Medicine, 379(2). 10.1056/nejmoa1804710

6. Cardoso, F., van’t Veer, L. J., Bogaerts, J., Slaets, L., Viale, G., Delaloge, S., Pierga, J.-Y., Brain, E., Causeret, S., DeLorenzi, M., Glas, A. M., Golfinopoulos, V., Goulioti, T., Knox, S., Matos, E., Meulemans, B., Neijenhuis, P. A., Nitz, U., Passalacqua, R., … Piccart, M. (2016). 70-Gene Signature as an Aid to Treatment Decisions in Early-Stage Breast Cancer. New England Journal of Medicine, 375(8). 10.1056/nejmoa1602253

7. Piccart, M., van ’t Veer, L. J., Poncet, C., Lopes Cardozo, J. M. N., Delaloge, S., Pierga, J. Y., Vuylsteke, P., Brain, E., Vrijaldenhoven, S., Neijenhuis, P. A., Causeret, S., Smilde, T. J., Viale, G., Glas, A. M., Delorenzi, M., Sotiriou, C., Rubio, I. T., Kümmel, S., Zoppoli, G., … Rutgers, E. J. T. (2021). 70-gene signature as an aid for treatment decisions in early breast cancer: updated results of the phase 3 randomised MINDACT trial with an exploratory analysis by age. The Lancet Oncology, 22(4). 10.1016/S1470-2045(21)00007-3

8. Torossian, N., Gabriel, M., Papoutsoglou, P., Foretek, D., Brochard, C., Kamal, M., Ramdani, L., Lamy, C., Lecerf, C., Halladjian, M., Dupain, C., Lafleur, J., Aguilar-Mahecha, A., Basik, M., Vincent-Salomon, A., Tourneau, C. L. E., Roman-Roman, S., Gautheret, D., & Morillon, A. (2025). Reference-free RNA profiling predicts triple negative breast cancer chemoresistance to neoadjuvant treatment. NAR Cancer, 7(4). 10.1093/narcan/zcaf036

9. Freitas, A. J. A., Nunes, C. R., Mano, M. S., Causin, R. L., Santana, I. V. V., de Oliveira, M. A., Calfa, S., Silveira, H. C. S., de Pádua Souza, C., & Marques, M. M. C. (2023). Gene expression alterations predict the pathological complete response in triple-negative breast cancer exploratory analysis of the NACATRINE trial. Scientific Reports, 13(1). 10.1038/s41598-023-48657-6

10. Omar, M., Nuzzo, P. V., Ravera, F., Bleve, S., Fanelli, G. N., Zanettini, C., Valencia, I., & Marchionni, L. (2023). Notch-based gene signature for predicting the response to neoadjuvant chemotherapy in triple-negative breast cancer. Journal of Translational Medicine, 21(1). 10.1186/s12967-023-04713-3

11. Adam, G., Rampášek, L., Safikhani, Z., Smirnov, P., Haibe-Kains, B., & Goldenberg, A. (2020). Machine learning approaches to drug response prediction: challenges and recent progress. Npj Precision Oncology 2020 4:1, 4(1), 1–10. 10.1038/s41698-020-0122-1

12. Sammut, S. J., Crispin-Ortuzar, M., Chin, S. F., Provenzano, E., Bardwell, H. A., Ma, W., Cope, W., Dariush, A., Dawson, S. J., Abraham, J. E., Dunn, J., Hiller, L., Thomas, J., Cameron, D. A., Bartlett, J. M. S., Hayward, L., Pharoah, P. D., Markowetz, F., Rueda, O. M., … Caldas, C. (2021). Multi-omic machine learning predictor of breast cancer therapy response. Nature 2021 601:7894, 601(7894), 623–629. 10.1038/s41586-021-04278-5

13. Yuan, M., Liu, H., Huang, Y. E., Hou, F., Wang, L., Wang, Q., & Jiang, W. (2025). HAPIR: a refined Hallmark gene set-based machine learning approach for predicting immunotherapy response in cancer patients. Npj Precision Oncology, 9(1). 10.1038/s41698-025-00992-9

14. Santos, C. S., & Amorim-Lopes, M. (2025). Externally validated and clinically useful machine learning algorithms to support patient-related decision-making in oncology: a scoping review. BMC Medical Research Methodology, 25(1). 10.1186/s12874-025-02463-y

15. Zhang, Z., Hernandez, K., Savage, J., Li, S., Miller, D., Agrawal, S., Ortuno, F., Staudt, L. M., Heath, A., & Grossman, R. L. (2021). Uniform genomic data analysis in the NCI Genomic Data Commons. Nature Communications, 12(1). 10.1038/s41467-021-21254-9

16. Bernard, P. S., Parker, J. S., Mullins, M., Cheung, M. C. U., Leung, S., Voduc, D., Vickery, T., Davies, S., Fauron, C., He, X., Hu, Z., Quackenhush, J. F., Stijleman, I. J., Palazzo, J., Matron, J. S., Nobel, A. B., Mardis, E., Nielsen, T. O., Ellis, M. J., & Perou, C. M. (2009). Supervised risk predictor of breast cancer based on intrinsic subtypes. Journal of Clinical Oncology, 27(8). 10.1200/JCO.2008.18.1370

17. Kuenzi, B. M., Park, J., Fong, S. H., Sanchez, K. S., Lee, J., Kreisberg, J. F., Ma, J., & Ideker, T. (2020). Predicting Drug Response and Synergy Using a Deep Learning Model of Human Cancer Cells. Cancer Cell, 38(5), 672–684.e6. 10.1016/J.CCELL.2020.09.014

18. Sharifi-Noghabi, H., Zolotareva, O., Collins, C. C., & Ester, M. (2019). MOLI: Multi-omics late integration with deep neural networks for drug response prediction. Bioinformatics, 35(14). 10.1093/bioinformatics/btz318

19. Basu, A., Mitra, R., Liu, H., Schreiber, S. L., & Clemons, P. A. (2018). RWEN: Response-weighted elastic net for prediction of chemosensitivity of cancer cell lines. Bioinformatics, 34(19). 10.1093/bioinformatics/bty199

20. Geeleher, P., Cox, N., & Stephanie Huang, R. (2014). PRRophetic: An r package for prediction of clinical chemotherapeutic response from tumor gene expression levels. PLoS ONE, 9(9). 10.1371/journal.pone.0107468

21. Guan, N. N., Zhao, Y., Wang, C. C., Li, J. Q., Chen, X., & Piao, X. (2019). Anticancer Drug Response Prediction in Cell Lines Using Weighted Graph Regularized Matrix Factorization. Molecular Therapy Nucleic Acids, 17. 10.1016/j.omtn.2019.05.017

22. Albain, K., Anderson, S., Arriagada, R., Barlow, W., Bergh, J., Bliss, J., Buyse, M., Cameron, D., Carrasco, E., Clarke, M., Correa, C., Coates, A., Collins, R., Costantino, J., Cutter, D., Cuzick, J., Darby, S., Davidson, N., Davies, C., … Wood, W. (2012). Comparisons between different polychemotherapy regimens for early breast cancer: Meta-analyses of long-term outcome among 100 000 women in 123 randomised trials. The Lancet, 379(9814). 10.1016/S0140-6736(11)61625-5

23. Bear, H. D., Anderson, S., Brown, A., Smith, R., Mamounas, E. P., Fisher, B., Margolese, R., Theoret, H., Soran, A., Lawrence Wickerham, D., & Wolmark, N. (2003). The effect on tumor response of adding sequential preoperative docetaxel to preoperative doxorubicin and cyclophosphamide: Preliminary results from National Surgical Adjuvant Breast and Bowel Project Protocol B-27. Journal of Clinical Oncology, 21(22). 10.1200/JCO.2003.12.005

24. von Minckwitz, G., Untch, M., Blohmer, J. U., Costa, S. D., Eidtmann, H., Fasching, P. A., Gerber, B., Eiermann, W., Hilfrich, J., Huober, J., Jackisch, C., Kaufmann, M., Konecny, G. E., Denkert, C., Nekljudova, V., Mehta, K., & Loibl, S. (2012). Definition and impact of pathologic complete response on prognosis after neoadjuvant chemotherapy in various intrinsic breast cancer subtypes. Journal of Clinical Oncology, 30(15). 10.1200/JCO.2011.38.8595

25. Ellis, M. J., Coop, A., Singh, B., Mauriac, L., Llombert-Cussac, A., Jänicke, F., Miller, W. R., Evans, D. B., Dugan, M., Brady, C., Quebe-Fehling, E., & Borgs, M. (2001). Letrozole is more effective neoadjuvant endocrine therapy than tamoxifen for ErbB-1-and/or ErbB-2-positive, estrogen receptor-positive primary breast cancer: Evidence from a phase III randomized trial. Journal of Clinical Oncology, 19(18). 10.1200/JCO.2001.19.18.3808

26. Smith, I. E., Dowsett, M., Ebbs, S. R., Dixon, J. M., Skene, A., Blohmer, J. U., Ashley, S. E., Francis, S., Boeddinghaus, I., & Walsh, G. (2005). Neoadjuvant treatment of postmenopausal breast cancer with anastrozole, tamoxifen, or both in combination: The Immediate Preoperative Anastrozole, Tamoxifen, or Combined With Tamoxifen (IMPACT) multicenter double-blind randomized trial. Journal of Clinical Oncology, 23(22). 10.1200/JCO.2005.04.005

27. Bonneterre, J., Thürlimann, B., Robertson, J. F. R., Krzakowski, M., Mauriac, L., Koralewski, P., Vergote, I., Webster, A., Steinberg, M., & von Euler, M. (2000). Anastrozole versus tamoxifen as first-line therapy for advanced breast cancer in 668 postmenopausal women: Results of the tamoxifen or arimidex randomized group efficacy and tolerability study. Journal of Clinical Oncology, 18(22). 10.1200/JCO.2000.18.22.3748

28. Paridaens, R. J., Dirix, L. Y., Beex, L. v., Nooij, M., Cameron, D. A., Cufer, T., Piccart, M. J., Bogaerts, J., & Therasse, P. (2008). Phase III study comparing exemestane with tamoxifen as first-line hormonal treatment of metastatic breast cancer in postmenopausal women: The European Organisation for Research and Treatment of Cancer Breast Cancer Cooperative Group. Journal of Clinical Oncology, 26(30). 10.1200/JCO.2007.14.4659

29. Muss, H. B., Wells, H. B., Paschold, E. H., Black, W. R., Cooper, M. R., Capizzi, R. L., Christian, R., Cruz, J. M., Jackson, D. v., & Powell, B. L. (1988). Megestrol acetate versus tamoxifen in advanced breast cancer: 5-year analysis--a phase III trial of the Piedmont Oncology Association. Journal of Clinical OncologyJ: Official Journal of the American Society of Clinical Oncology, 6(7). 10.1200/JCO.1988.6.7.1098

30. Slamon, D. J., Clark, G. M., Wong, S. G., Levin, W. J., Ullrich, A., & McGuire, W. L. (1987). Human breast cancer: Correlation of relapse and survival with amplification of the HER-2/neu oncogene. Science, 235(4785). 10.1126/science.3798106

31. Vogel, C. L., Cobleigh, M. A., Tripathy, D., Gutheil, J. C., Harris, L. N., Fehrenbacher, L., Slamon, D. J., Murphy, M., Novotny, W. F., Burchmore, M., Shak, S., Stewart, S. J., & Press, M. (2023). Efficacy and Safety of Trastuzumab as a Single Agent in First-Line Treatment of HER2-Overexpressing Metastatic Breast Cancer. Journal of Clinical Oncology, 41(9). 10.1200/JCO.22.02516

32. Baselga, J., Carbonell, X., Castañeda-Soto, N. J., Clemens, M., Green, M., Harvey, V., Morales, S., Barton, C., & Ghahramani, P. (2005). Phase II study of efficacy, safety, and pharmacokinetics of trastuzumab monotherapy administered on a 3-weekly schedule. Journal of Clinical Oncology, 23(10). 10.1200/JCO.2005.01.014

33. Buzdar, A. U., Ibrahim, N. K., Francis, D., Booser, D. J., Thomas, E. S., Theriault, R. L., Pusztai, L., Green, M. C., Arun, B. K., Giordano, S. H., Cristofanilli, M., Frye, D. K., Smith, T. L., Hunt, K. K., Singletary, S. E., Sahin, A. A., Ewer, M. S., Buchholz, T. A., Berry, D., & Hortobagyi, G. N. (2005). Significantly higher pathologic complete remission rate after neoadjuvant therapy with trastuzumab, paclitaxel, and epirubicin chemotherapy: Results of a randomized trial in human epidermal growth factor receptor 2-positive operable breast cancer. Journal of Clinical Oncology, 23(16). 10.1200/JCO.2005.07.032

34. Gianni, L., Eiermann, W., Semiglazov, V., Lluch, A., Tjulandin, S., Zambetti, M., Moliterni, A., Vazquez, F., Byakhov, M. J., Lichinitser, M., Climent, M. A., Ciruelos, E., Ojeda, B., Mansutti, M., Bozhok, A., Magazzù, D., Heinzmann, D., Steinseifer, J., Valagussa, P., & Baselga, J. (2014). Neoadjuvant and adjuvant trastuzumab in patients with HER2-positive locally advanced breast cancer (NOAH): Follow-up of a randomised controlled superiority trial with a parallel HER2-negative cohort. The Lancet Oncology, 15(6). 10.1016/S1470-2045(14)70080-4

35. Untch, M., Rezai, M., Loibl, S., Fasching, P. A., Huober, J., Tesch, H., Bauerfeind, I., Hilfrich, J., Eidtmann, H., Gerber, B., Hanusch, C., Kühn, T., du Bois, A., Blohmer, J. U., Thomssen, C., Dan Costa, S., Jackisch, C., Kaufmann, M., Mehta, K., & von Minckwitz, G. (2010). Neoadjuvant treatment with trastuzumab in HER2-positive breast cancer: Results from the GeparQuattro study. Journal of Clinical Oncology, 28(12). 10.1200/JCO.2009.23.8451

